# Impact of the COVID-19 pandemic on food safety inspection outcomes in Toronto, Canada: a Bayesian interrupted time series analysis

**DOI:** 10.1101/2023.01.06.23284283

**Authors:** Ian Young, Binyam Negussie Desta, Fatih Sekercioglu

## Abstract

The coronavirus disease (COVID-19) pandemic resulted in major disruptions to the food service industry and regulatory food inspections. The objective of this study was to conduct an interrupted time series analysis to investigate the impact of the COVID-19 pandemic on food safety inspection trends in Toronto, Canada. Inspection data for restaurants and take-out establishments were obtained from 2017 to 2022 and ordered as a weekly time series. Bayesian segmented regression was conducted to evaluate the impact of the pandemic on weekly infraction and inspection pass rates. On average, a 0.31-point lower weekly infraction rate (95% credible interval [CI]: 0.23, 0.40) and a 2.0% higher probability of passing inspections (95% CI: 1.1%, 3.0%) were predicted in the pandemic period compared to pre-pandemic. Models predicted lower infraction rates and higher pass rates immediately following the pandemic that were regressing back toward pre-pandemic levels in 2022. Seasonal effects were also identified, with infraction rates highest in April and pass rates lowest in August. The COVID-19 pandemic resulted in an initial positive effect on food safety outcomes in restaurants and take-out food establishments in Toronto, but this effect appears to be temporary. Additional research is needed on seasonal and long-term inspection trends post-pandemic.

## Introduction

The coronavirus disease 2019 (COVID-19) pandemic caused major disruptions to society when declared by the World Health Organization in March 2020. Jurisdictions across the world instituted lockdowns, physical distancing requirements, and temporary closures of public businesses and other facilities to control the spread of the virus [1]. During this time, the incidence of food-borne illness reported to the United States’ Foodborne Diseases Active Surveillance Network was 26% lower than in the prior three years (2017-2019) [2]. Similarly, in Colorado, a 52% decrease in the rate of persons seeking medical care for acute gastroenteritis was observed in 2020 vs. 2017-2019 [3]. It is not clear to what extent these decreases were due to actual declines in food-borne illness exposures or to decreased illness reporting, detection, or diagnosis.

Restaurants and food service settings were intermittently closed to indoor dining during the initial and subsequent waves of the COVID-19 pandemic, and as a result, they shifted their food service operations primarily to take-out and delivery, curbside pickup, and drive-thru options during pandemic waves [4]. The pandemic also led to enhanced hygiene, cleaning, disinfection, and other infection control practices and policies at these establishments as they aimed to control the spread of the virus (e.g., increased hand hygiene, cleaning and disinfection of high-touch surfaces, contactless menus and payment options, physical distancing of staff and patrons, suspending of self-serve buffets and salad bars) [5]. These enhanced practices and policies could also have influenced a reduction in food contamination and the spread of foodborne pathogens via these settings, which are common sources of food-borne illness outbreaks [6]. However, little research has been conducted to investigate food safety practices at restaurants and other food service establishments during the COVID-19 pandemic.

In Toronto, Canada, the city’s public health inspectors are responsible for conducting routine food safety inspections of restaurants and other retail food establishments according to provincial guidelines [7]. Food establishments are inspected once, twice, or three times per year depending on their risk level (low, moderate, or high, respectively) as determined via a risk categorization process and in accordance with the provincial food safety regulation [7]. In Toronto, food establishments then receive a pass, conditional pass, or fail rating based on the inspection results as part of a public disclosure system called DineSafe [8]. The inspection rating must be visible from the establishments entrance and results are posted publicly online [8]. Routine food safety inspections are important to promote and encourage food safety practices, and they can serve as an early warning indicator of the potential for food-borne illness outbreaks [9–11]. The pandemic led to an initial pause in food safety inspections in Toronto immediately following the provincial emergency declaration in March 2020, but its impacts on inspection outcomes have not been investigated. We conducted an interrupted time series analysis to investigate the impact of the COVID-19 pandemic on DineSafe inspection trends in Toronto. Results can inform public health policy and planning related to food inspections in the post-pandemic era and in future pandemic preparedness.

## Materials and Methods

### Dataset access and description

We obtained DineSafe inspection data from the City of Toronto’s Open Data Portal [12]. As the open data repository only provides the most recent two years of inspection data, we made a special request to access data dating back to 2017. Our dataset timeframe ranged form 1 January 2017 to 31 December 2022. The dataset contained information on the establishment ID number, name, business type, address, geolocation, and inspections results. Inspection results included a separate row for each infraction identified, including its severity (minor, significant, crucial, or other) and description, as well as the overall inspection rating (pass, conditional pass, or fail). The full dataset contained information on 218,607 total inspection outcomes.

### Dataset preparation

Data were obtained as comma separated value files and imported into RStudio (version 2022.12.0 running R 4.2.2) for formatting, preparation, and analysis [13,14]. For the purposes of this analysis, we restricted the dataset to establishments classified as restaurants and food take-aways. We then reformatted the dataset to one row per inspection by summing the number of infractions per inspection. As conditional pass and fail inspections usually resulted in a re-inspection within 48 hours, we removed such re-inspections from the dataset to focus only on the routine inspections. The dataset was then converted to a time series for analysis. We created a weekly time variable and summarized the total number of inspections conducted, pass ratings, and infractions identified each week.

In Ontario, the province declared a state of emergency for COVID-19 on 17 March 2020, resulting in the first provincial lockdown. Starting on this date, the DineSafe program was essentially paused for several weeks. The dataset contained zero to three inspections per week from week 13 to week 25 in 2020 (23 March to 21 June), which likely reflected only complaint-based inspections. Given the low and inconsistent numbers of inspections during these weeks, we decided to remove these weeks from the dataset. Therefore, the final dataset for analysis contained 297 weeks of data: 168 weeks prior to the pandemic, and 129 weeks during the pandemic.

### Interrupted time series analysis

We conducted an interrupted time series analysis using segmented regression to evaluate the impact of the pandemic on infraction and pass rate outcomes [15,16]. Analysis was conducted under a Bayesian framework [17]. Bayesian analysis incorporates uncertainty via estimation of posterior distributions of model parameters, and direct probability statements can be made based on model results [17,18]. Two models were constructed to assess each of two outcomes of interest: 1) a negative binomial regression model for weekly infraction counts, with an offset term to account for the number of inspections conducted; and 2) an aggregated logistic regression model for the number of weekly pass ratings (vs. conditional pass or fail ratings) per number of inspections conducted [17].

The following variables were included as fixed effects in each model: time elapsed (continuous), pandemic period (yes/no indicator variable), and an interaction term between these two variables [15,16]. The pandemic period was defined to start the week following the first lockdown and emergency declaration in Ontario (Mar. 23, 2020). To account for seasonal effects, we included month as a varying effect via a multi-level structure. For each outcome, we compared this model structure to a model with month as a fixed-effect indicator variable. We also compared both models to models that included a first-order autoregressive term to account for any residual autocorrelation in weekly observations. These model comparisons were conducted using leave-one-out (LOO) cross-validation [19].

Weekly informative priors (prior distributions) were specified for all model beta coefficient parameters [18,20]. Priors were specified to have normal distributions with a mean of 0 and standard deviation (SD) of 1. Priors for varying effects (month) and residual SD parameters used half Student-*t* priors with 3 degrees of freedom and a SD of 2.5 [20]. The appropriateness of these priors was assessed via prior predictive checking to ensure the prior distributions captured a reasonable range of plausible values. Sensitivity analysis was also conducted to assess the impact of specifying alternative weakly informative priors for beta coefficient parameters of *N*(0,0.5) and *N*(0,2).

Models were constructed using the “brms” package in RStudio, which fits models via the probabilistic programming software Stan [20,21]. We used the “CmdStanR” interface to fit the Stan models [22]. Stan estimates model parameters using Hamiltonian Monte Carlo and its extension, the no-U-turn sampler [20,21]. We estimated models using 4000 iterations across each of four chains via four cores, of which the first 1000 were warmup iterations. Model convergence was assessed via examination of trace plots, effective sample sizes, and r-hat values [17,20,23]. We also conducted posterior predictive checks to evaluate the suitability of the models to simulate new data in relation to the observed data, and evaluated residual autocorrelation for model parameters [24].

To facilitate and visualize interpretation of the model parameters, we calculated posterior predictions of the expected value of model parameters using the “marginaleffects” package [25]. Average marginal effects (i.e., contrasts) were calculated to assess the effect of the pandemic on both outcomes. Long-term time trends and the conditional seasonal effect of month were also visualized in the pre-pandemic and pandemic periods. All figures show the parameter distribution densities along with the median value and 66% and 95% credible intervals (CI). A copy of the dataset used in this analysis along with R script files used for all formatting and analysis in this study are available from the following GitHub page: https://github.com/iany33/dinesafepandemic.

## Results

### Descriptive results

The time series dataset contained data on a total of 81,435 inspections. The overall inspection pass rate was 91.4% (74,408 / 81,435). A total of 103,118 infractions were identified. Figure 1 shows weekly infraction and pass rates across the study time period, while Figure 2 shows the total number of infractions identified and inspections conducted per week. Raw summary comparisons for all outcomes identified before and during the pandemic are shown in Table 1. Weekly infraction rates appeared to be lower during compared to prior to the pandemic, while pass rates appeared similar (Figure 1 and Table 1). Additional variability was also noted for both outcomes during the pandemic period. The number of weekly inspections conducted was much lower during the pandemic period and started to increase back toward pre-pandemic levels in the latter part of 2022 (Figure 2 and Table 1).

**Table 1.**
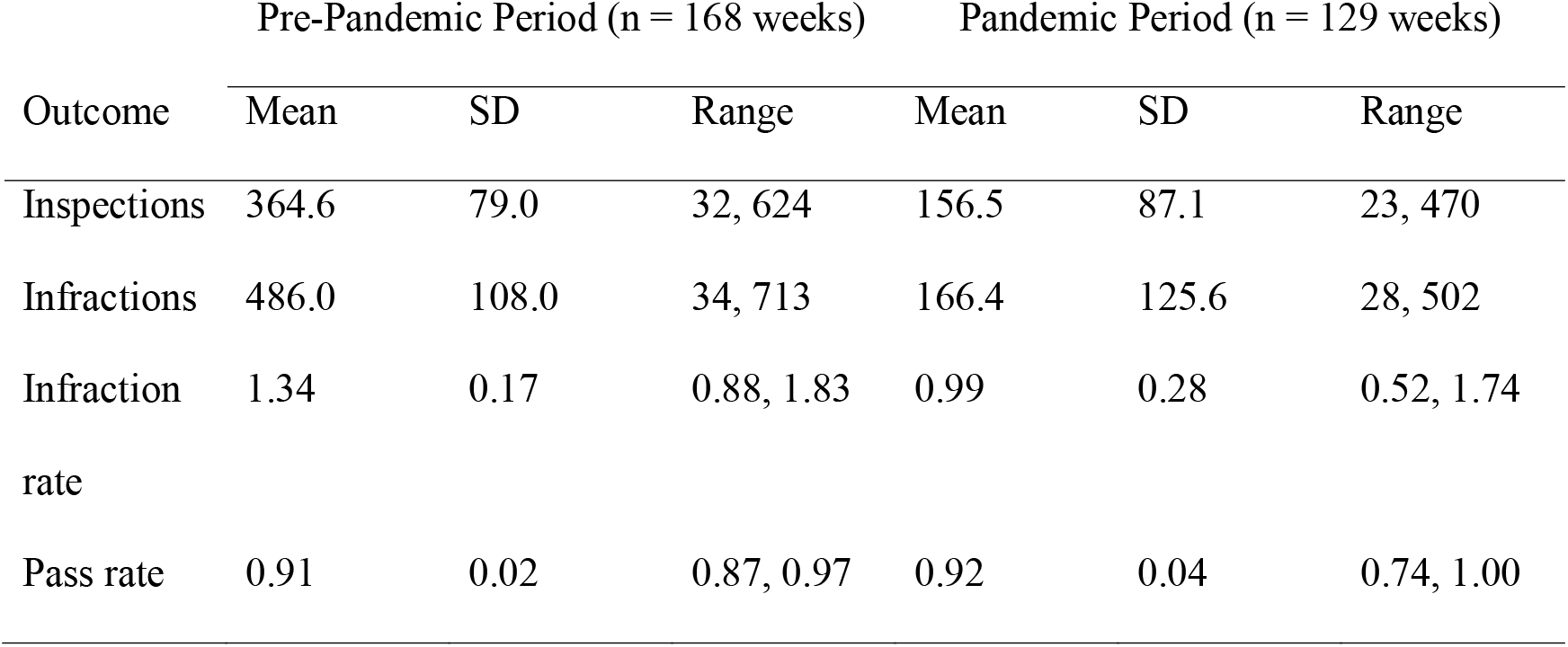
Summary statistics of weekly restaurant and take-away establishment food safety inspection trends, Toronto, 2017–2022.

| Outcome | Pre-Pandemic Period (n = 168 weeks) |  |  | Pandemic Period (n = 129 weeks) |  |  |
| --- | --- | --- | --- | --- | --- | --- |
|  | Mean | SD | Range | Mean | SD | Range |
| Inspections | 364.6 | 79.0 | 32, 624 | 156.5 | 87.1 | 23, 470 |
| Infractions | 486.0 | 108.0 | 34, 713 | 166.4 | 125.6 | 28, 502 |
| Infraction rate | 1.34 | 0.17 | 0.88, 1.83 | 0.99 | 0.28 | 0.52, 1.74 |
| Pass rate | 0.91 | 0.02 | 0.87, 0.97 | 0.92 | 0.04 | 0.74, 1.00 |

**Figure 1.**
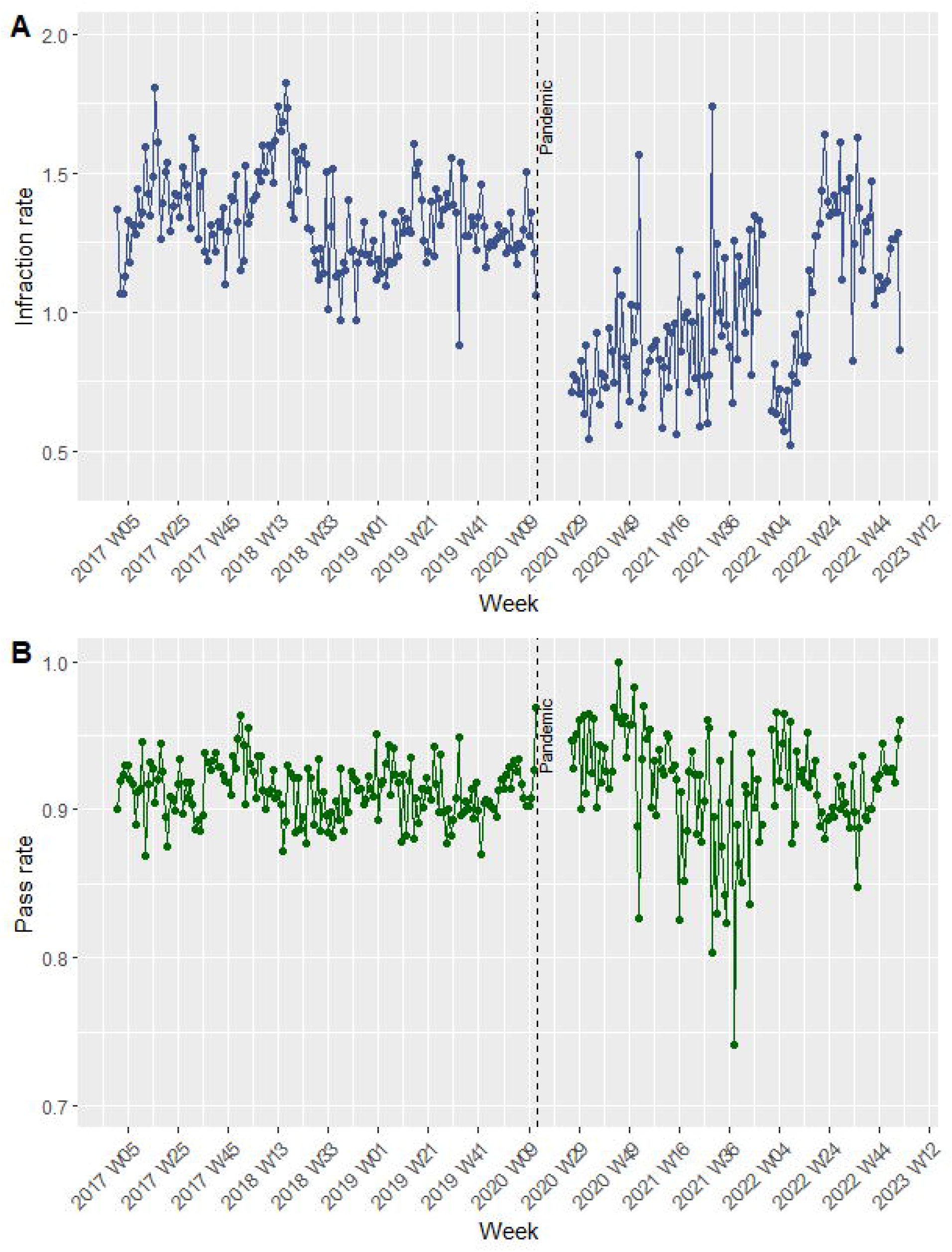
A) Weekly infraction rate (number of total infractions identified per number of inspections conducted) at restaurants and take-out establishments in Toronto, 2017–2022. B) Weekly pass rate (number of passes per number of inspections conducted) at restaurants and take-out establishments in Toronto, 2017–2022.

**Figure 2.**
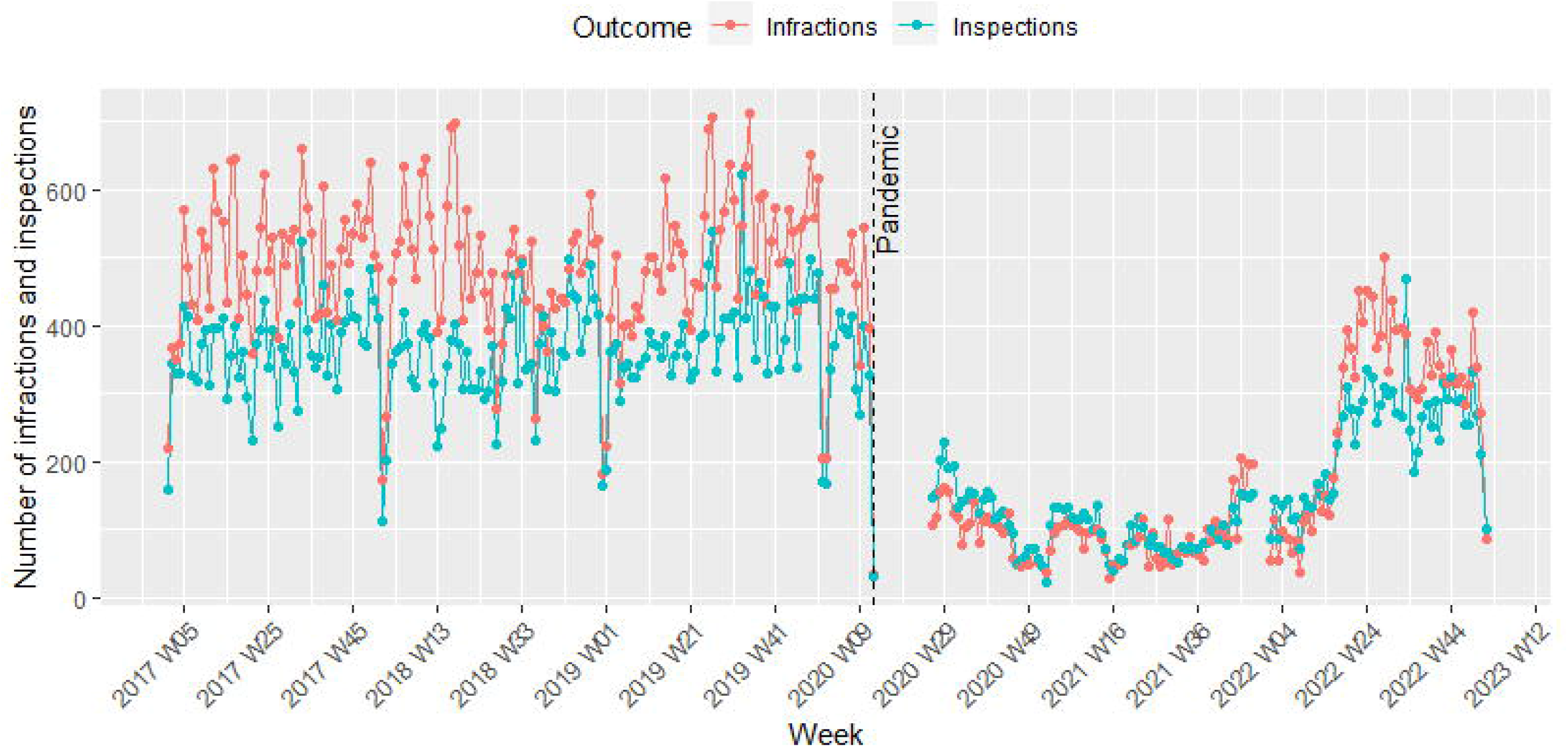
Number of total infractions identified and inspections conducted per week at restaurants and take-out establishments in Toronto, 2017–2022.

### Interrupted time series results

Both the infraction rate and pass rate models showed no issues with convergence or residual autocorrelation (see Supplementary Material). Additionally, for both outcomes, the LOO comparisons found that multi-level models with month as a varying effect, and no auto-regressive term, had the best fit (see Supplementary Material). The sensitivity analysis of alternative prior distributions suggested minimal impact on the model results (see Supplementary Material).

Results of the multi-level negative binomial regression model for weekly infraction rates are shown in Table 2, with model coefficients exponentiated as incidence rate ratios (IRR). However, the effects are most intuitively illustrated in Figures 3-5. Figure 3 shows posterior predictions for the average expected value of the infraction rate in pre-pandemic and pandemic periods on the left, with the average marginal effect (i.e., contrast) of the pandemic shown on the right. On average, the pandemic resulted in a 0.31-point lower food safety infraction rate (95% CI: 0.23, 0.40) compared to the pre-pandemic period (Figure 3). This effect was primarily noted in the first 2 years of the pandemic until the week of 26 April 2022 (−0.40, 95% CI: -0.32, -0.48), when weekly inspection numbers were still consistently <200, compared to the most recent period from 22 April to 31 December 2022 (−0.09, 95% CI: -0.01, -0.19). Figure 4 shows posterior predictions for the infraction rate over time in both periods. A slightly decreasing trend in weekly infraction rates was noted pre-pandemic. A level change was noted post-pandemic, with much lower infraction rates that are predicted to be increasing back toward pre-pandemic levels (Figure 4). Figure 5 shows the monthly, or seasonal, effects of the pandemic on posterior predictions for the weekly infraction rate. In both pre-pandemic and pandemic periods, predicted infraction rates were highest in April, and lowest in January and September (Figure 5).

**Table 2.**
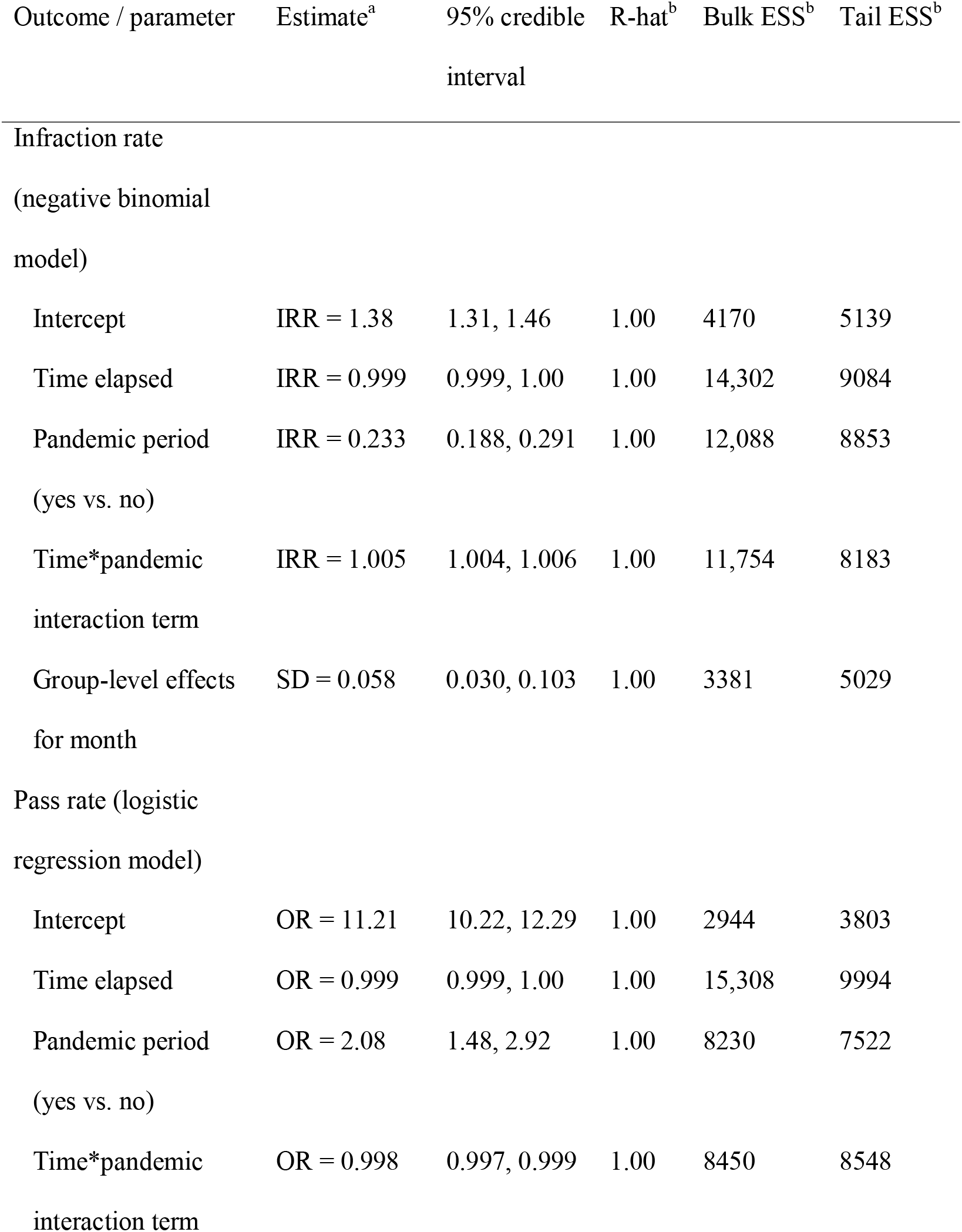
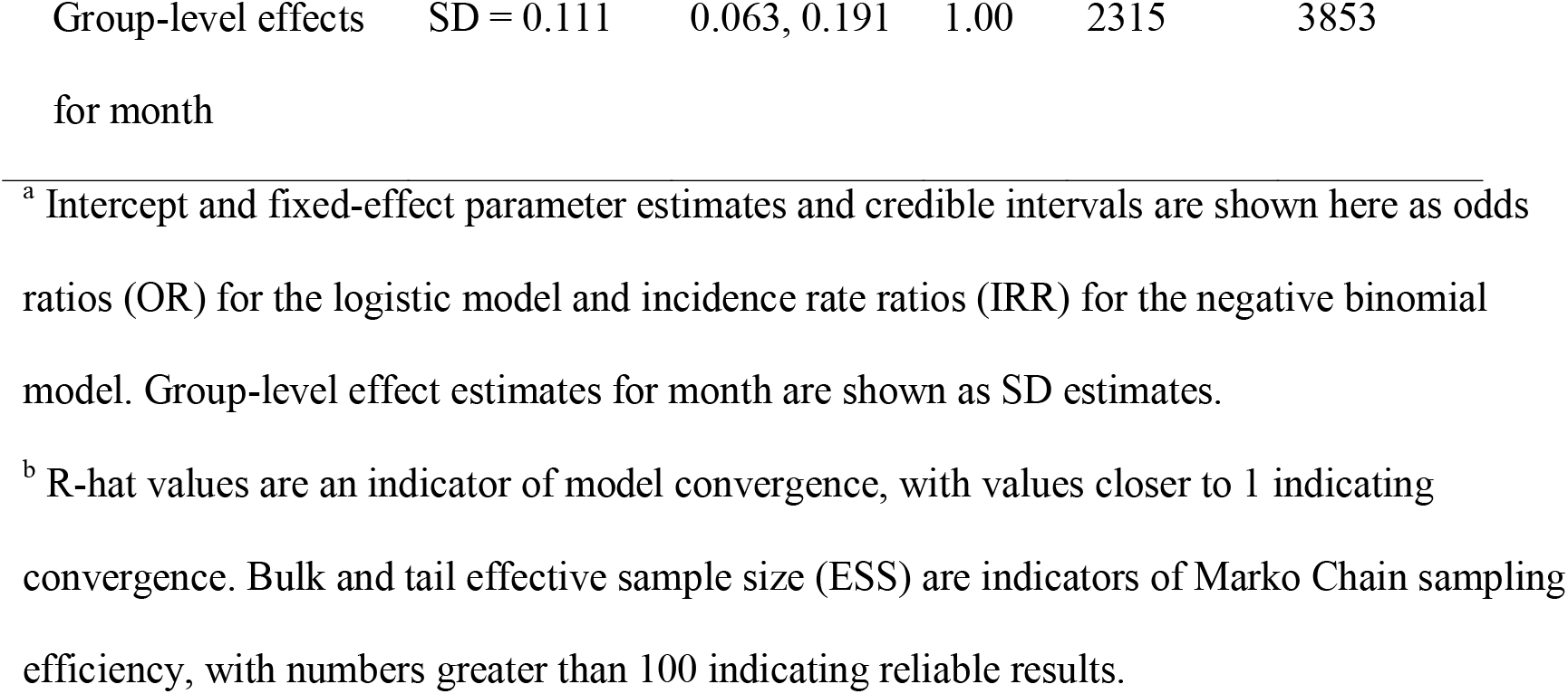
Bayesian segmented regression model results for two weekly inspection outcomes, Toronto, 2017–2022.

**Figure 3.**
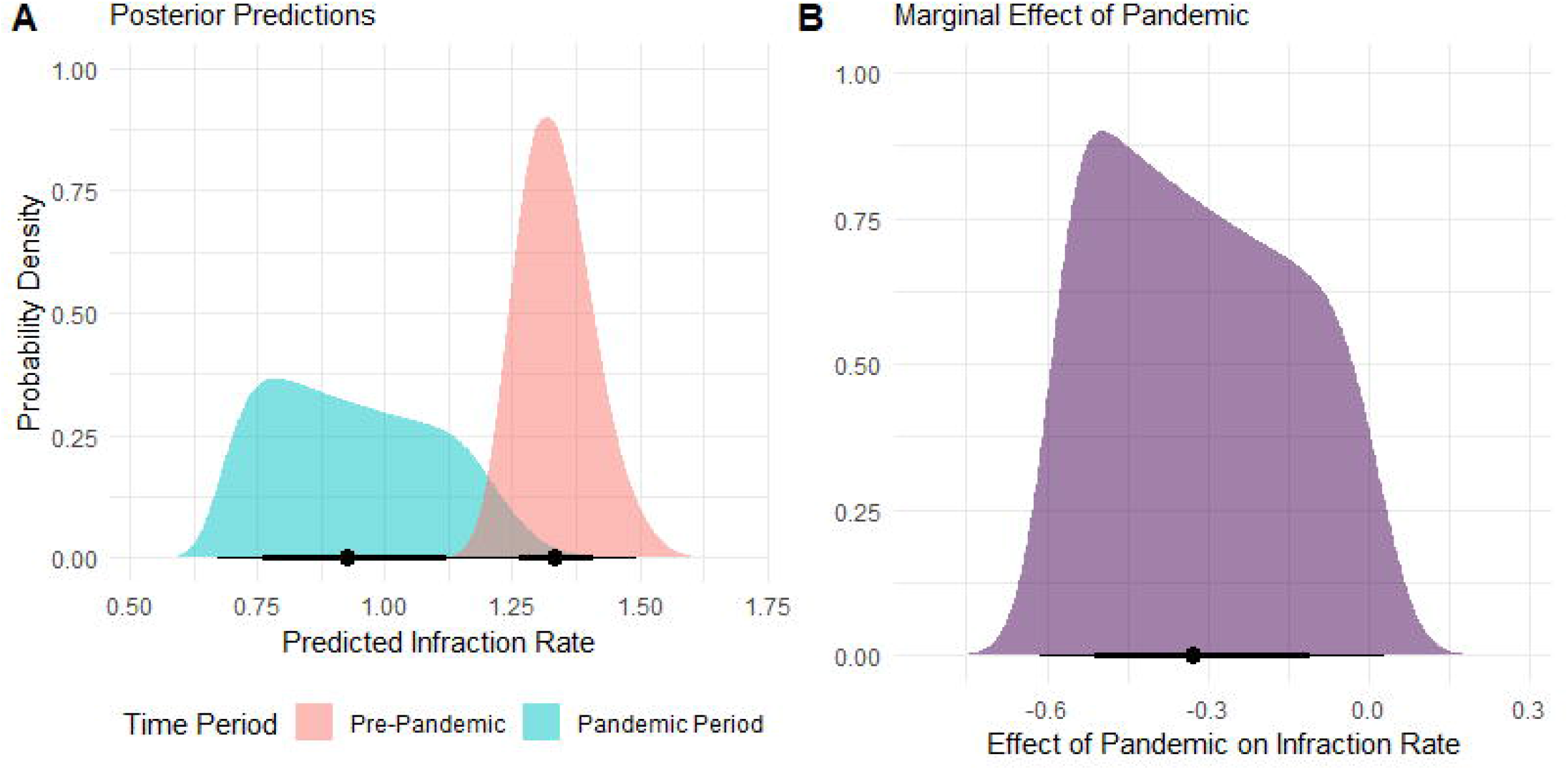
A) Posterior predictions of the average expected value of the weekly infraction rate at restaurants and take-out establishments in Toronto in the pre-pandemic (January 2017 to March 2020) and pandemic periods (June 2020 to December 2022). B) Average marginal effect of the COVID-19 pandemic on the expected value of the weekly infraction rate.

**Figure 4.**
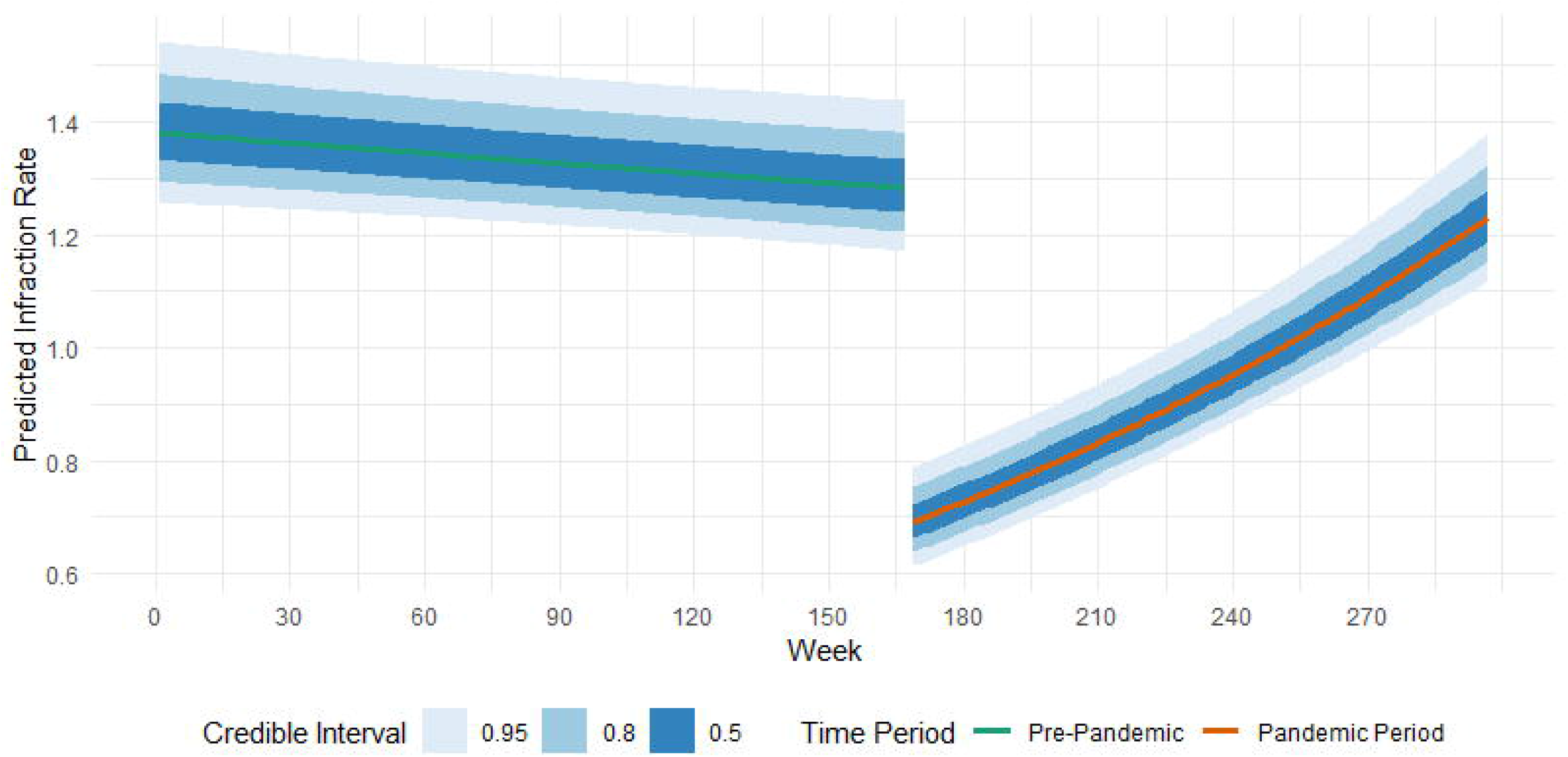
Predicted expected value of the weekly infraction rate at restaurants and take-out establishments in Toronto over time during the pre-pandemic (January 2017 to March 2020) and pandemic periods (June 2020 to December 2022).

**Figure 5.**
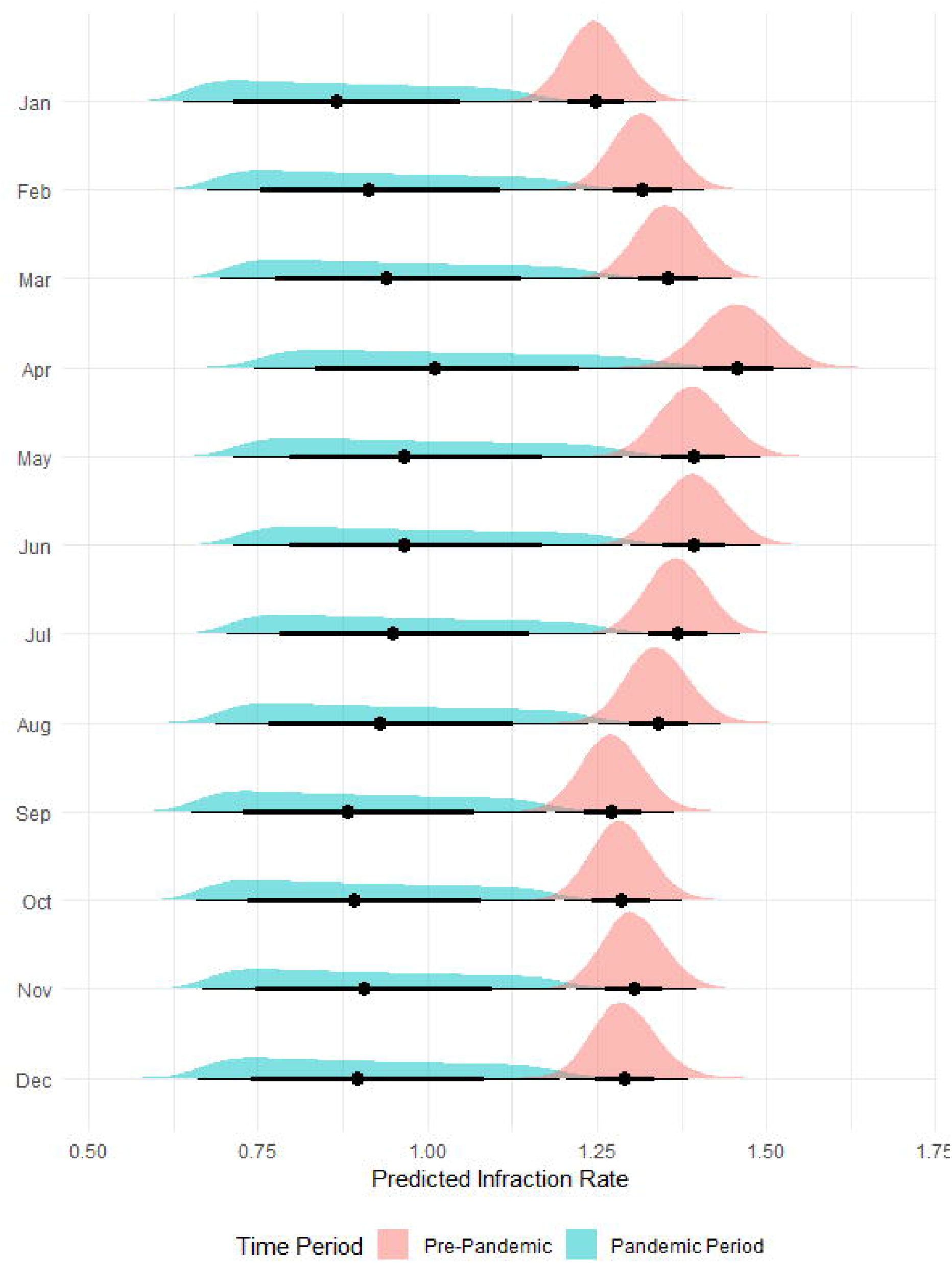
Posterior predictions of the month-specific average expected value of the weekly infraction rate at restaurants and take-out establishments in Toronto in the pre-pandemic (January 2017 to March 2020) and pandemic periods (June 2020 to December 2022).

Results of the multi-level aggregated logistic regression model for weekly pass rates are shown in Table 2, with model coefficients exponentiated as odds ratios (OR). As above, these effects are most intuitively illustrated in Figures 6-8. Figure 6 shows posterior predictions for the average expected value of the weekly inspection pass rate in pre-pandemic and pandemic periods on the left, with the average marginal effect of the pandemic shown on the right. On average, the pandemic resulted in a 2.0% higher probability of passing food safety inspections (95% CI: 1.1%, 3.0%) compared to the pre-pandemic period (Figure 6). As above, this effect was strongest in the first 2 years of the pandemic (2.3%, 95% CI: 1.4%, 3.2%) compared to the most recent period from 22 April to 31 December 2022 (1.4%, 95% CI: 0.3%, 2.6%). Figure 7 shows posterior predictions for the pass rate over time in both periods. A slightly decreasing trend in weekly infraction rates was noted pre-pandemic, but it is not clear if this is practically meaningful. A level change was noted after inspections were resumed during the pandemic, with much higher predicted pass rates that are regressing back toward pre-pandemic levels (Figure 7). Figure 8 shows the monthly, or seasonal, effects of the pandemic on posterior predictions for the weekly pass rate. In both pre-pandemic and pandemic periods, predicted pass rates were highest in January and December, and lowest in August (Figure 8).

**Figure 6.**
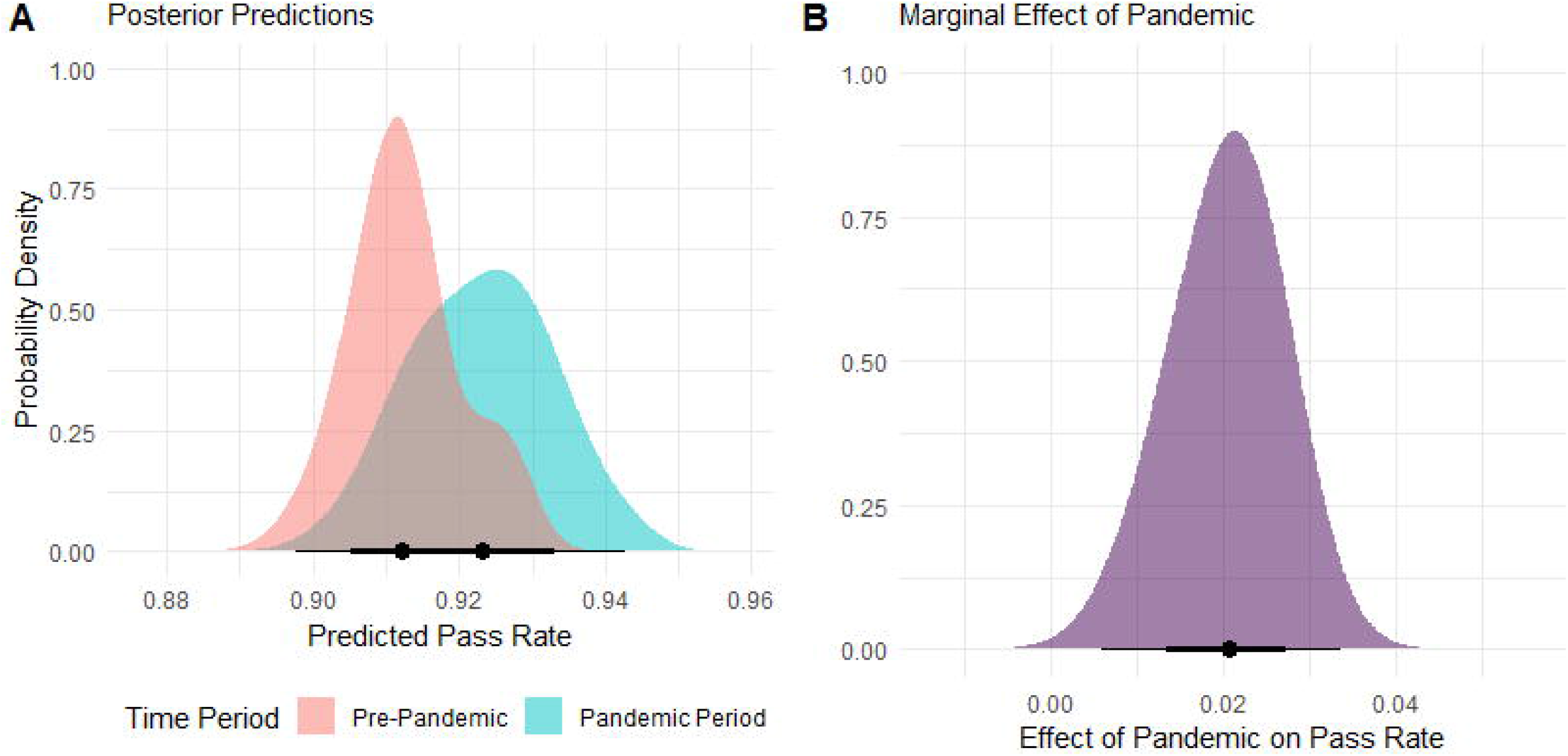
A) Posterior predictions of the average expected value of the weekly pass rate at restaurants and take-out establishments in Toronto in the pre-pandemic (January 2017 to March 2020) and pandemic periods (June 2020 to December 2022). B) Average marginal effect of the COVID-19 pandemic on the expected value of the weekly pass rate across the study time period.

**Figure 7.**
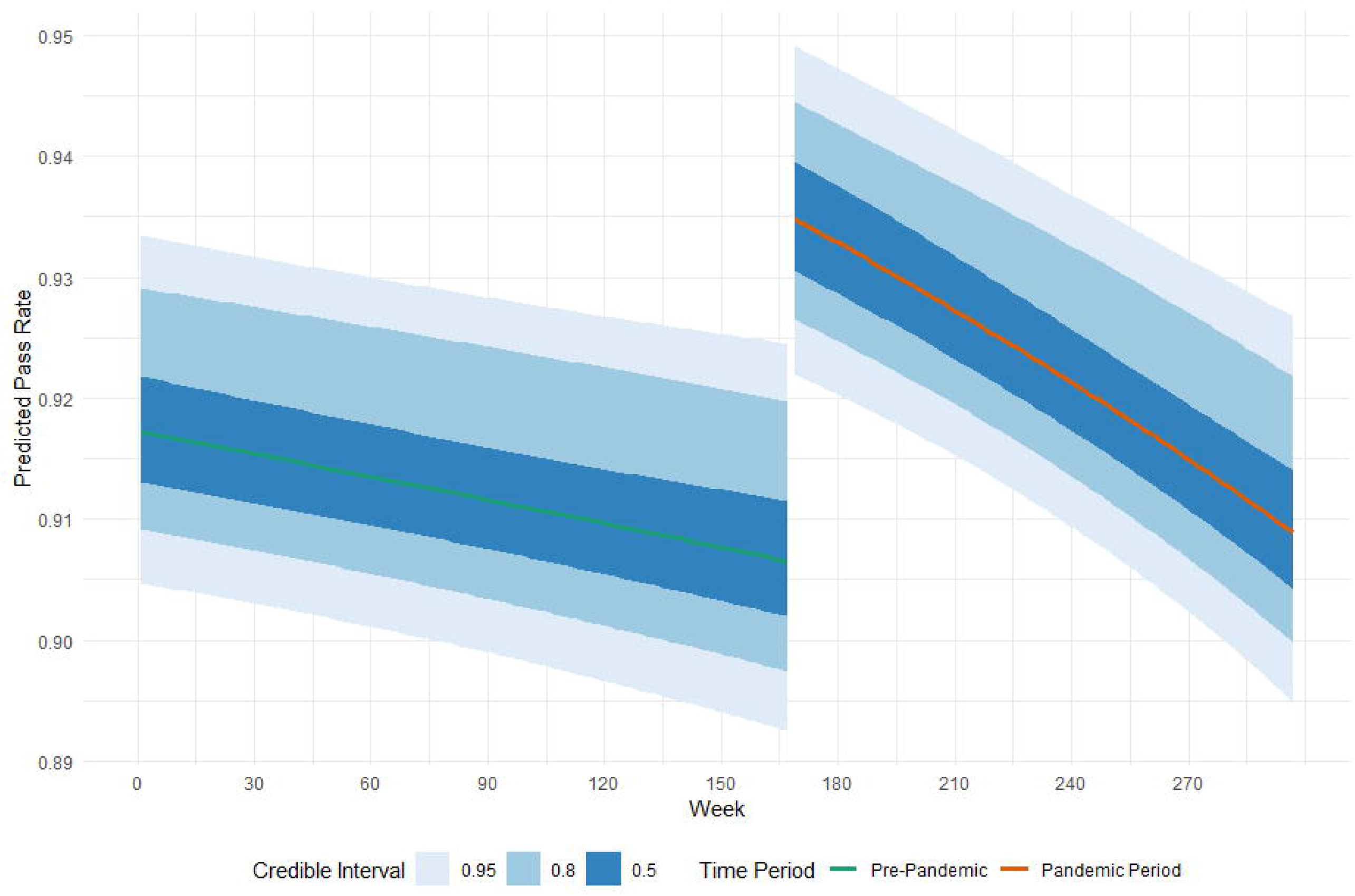
Predicted expected value of the weekly pass rate at restaurants and take-out establishments in Toronto over time during the pre-pandemic (January 2017 to March 2020) and pandemic periods (June 2020 to December 2022).

**Figure 8.**
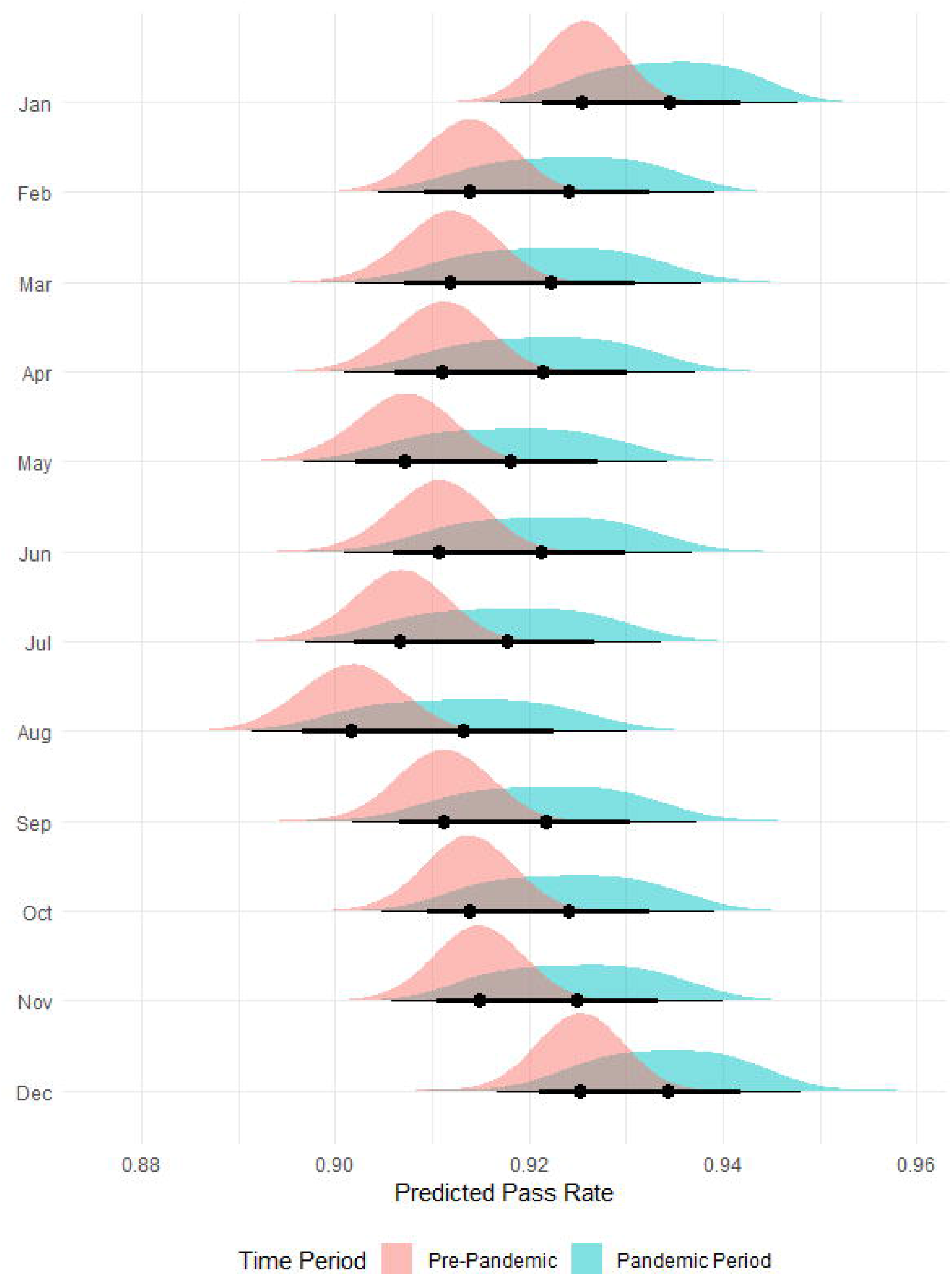
Posterior predictions of the month-specific average expected value of the weekly pass rate at restaurants and take-out establishments in Toronto in the pre-pandemic (January 2017 to March 2020) and pandemic periods (June 2020 to December 2022).

## Discussion

The COVID-19 pandemic resulted in major disruptions to society starting in March 2020, including routine public health services such as food safety inspections of restaurants and take-out facilities. This was reflected in the DineSafe program in Toronto, where inspections were not conducted for several weeks following the initial provincial declaration of emergency and lockdown. Additionally, the number of inspections conducted each week following resumption of routine inspections in 2020 was substantially reduced compared to pre-pandemic levels until approximately mid-2022. This reduction in DineSafe inspections is due to COVID-19 closures and restrictions, as well as the reallocation and secondment of many public health inspectors to assist with COVID-19 case and contact management and other pandemic-related duties during the initial and subsequent waves [26].

After controlling for long-term time trends and seasonality, our interrupted time series analysis found that there was a substantial average effect of the pandemic on lowering food safety infraction rates, and increasing inspection pass rates, particularly in the initial 1-2 years of the pandemic. This increase in food safety outcomes is likely due to the enhanced hygiene, cleaning, sanitation, and other precautions implemented in restaurants and take-out facilities to prevent the spread of the COVID-19 virus. Surveys and focus groups conducted with the general public in the initial months of the pandemic found an overall increased adoption of hand hygiene (e.g., handwashing, sanitizer use) and other cleaning and sanitation practices [27,28]. It is likely that these practices were also enhanced among restaurant and take-out facility management and staff due to personal concerns about COVID-19 and government requirements for infection control [29]. Additionally, food establishments were also likely responding to customer concerns about hygiene and COVID-19 control measures, which have been shown to be predictors of their dining behaviours at restaurants and other food service facilities during the pandemic [30].

Despite the average effects of the pandemic, when examining time trends during the pandemic period, we identified a regression of infraction and pass rates back toward pre-pandemic levels. The initial step change in rates following the pandemic and regression back toward the mean is common in interrupted time series analyses for temporary effects of major events or interventions [16]. It is unclear whether these rates will stabilize at or near pre-pandemic levels or whether they will worsen over time in the coming years. For example, the pandemic and other ongoing global events (e.g., inflation) have led to numerous operational challenges for restaurant and food service operators, including stress, financial losses, labour shortages, and supply chain difficulties that may persist in the future and lead to potential food safety lapses [31]. We also noted slightly decreasing trends for both rates during the pre-pandemic period, and it is unclear what was driving those trends or if they would have continued had the pandemic not occurred.

When examining infraction and pass rate trends by month, we identified some seasonal effects. The pre-pandemic and pandemic period monthly effects were similar, except there was much more variability noted in the pandemic period likely due to the lower number of inspections conducted and time trends noted above. Interestingly, weekly infraction rates were highest in the spring (April) and lowest in the fall to early winter (September to January), while inspection pass rates were lowest in the summer (August) and highest in winter (December and January). Conditional pass and fail outcomes for an inspection are assigned when one or more significant or critical infractions are identified that correspond to potential health hazards (e.g., time-temperature abuse, pest infestation) and cannot be immediately corrected [8]. In contrast, total infractions also include minor infractions that present minimal health risks directly (e.g., sanitary condition of food handling room) [8]. Lower pass rates in August could be related to higher average air temperatures in summer months. For example, prior research in New York has found that higher ambient air temperatures were associated with increased temperature-control related infractions in restaurants [32]. Foodborne illness rates also show a seasonal trend with many infections being highest in summer months [33,34]. It is unclear why total infractions differed seasonally. Future research would be beneficial in this area, including types of infractions identified and differences in their frequency by month or season, as well as primary research with public health inspectors and restaurant operators to determine barriers, facilitators, and other factors affecting food safety practices in these settings post-pandemic.

We examined total infraction rates as an outcome rather than specific categories (e.g., minor, significant, critical), as preliminary evaluation found little difference in trends when these rates were stratified by type, and the pass rate outcome reflects information on more severe infractions identified. The publicly available DineSafe dataset did not contain any information about the characteristics of included food establishments, such as inspection frequency, cuisine or food type, number of employees, or chain vs. independent status, which are known to be related to inspection outcomes but could not be investigated in this study [35–37]. Future research could aim to investigate how the pandemic affected food safety outcomes in food service establishments with different characteristics. The dataset contained geolocation data for each establishment, but these were not considered in this study as we focused on the overall time series of inspection outcomes across the city. Future research could also examine pandemic-related and post-pandemic inspection outcome trends in different geographical areas, as prior research has shown a relationship between infractions and neighbourhood socioeconomic status indicators [36,38].

We used a Bayesian approach to estimate the effect of the COVID-19 pandemic on food safety inspection outcomes in restaurants and take-out facilities in Toronto. This analytical approach allowed us to determine probability distributions of modelled outcomes and to account for uncertainty in the model parameters and predicted expected effects. We found that the pandemic had an initial positive effect on lowering total infraction rates and increasing inspection pass rates, but this effect appears to be temporary with outcomes regressing back toward pre-pandemic levels in 2022. This finding suggests that enhanced COVID-19 infection control measures could have temporarily improved food safety outcomes in restaurants and food service settings. However, further research is needed to examine longer-term trends in these outcomes as COVID-19 control measures and requirements are reduced and removed from such settings and as operators cope with additional post-pandemic stressors. We also identified seasonal trends in inspection outcomes that warrant future research and investigation.

## Data Availability

All data produced are available online at: https://github.com/iany33/dinesafepandemic

## Acknowledgements

The authors acknowledge the Open Data initiative at the City of Toronto for providing access to the DineSafe dataset for research and analysis purposes.

## References

1. Escandón K, et al. (2021) COVID-19 false dichotomies and a comprehensive review of the evidence regarding public health, COVID-19 symptomatology, SARS-CoV-2 transmission, mask wearing, and reinfection. BMC Infectious Diseases; 21: 710.

2. Ray LC, et al. (2021) Decreased incidence of infections caused by pathogens transmitted commonly through food during the COVID-19 pandemic — Foodborne Diseases Active Surveillance Network, 10 U.S. sites, 2017–2020. Morbidity and Mortality Weekly Report; 70: 1332–1336.

3. Armistead I, et al. (2022) Trends in outpatient medical-care seeking for acute gastroenteritis during the COVID-19 pandemic, 2020. Foodborne Pathogens and Disease; 19: 290–292.

4. Abebe GK, Charlebois S, Music J. (2022) Canadian consumers’ dining behaviors during the COVID-19 pandemic: implications for channel decisions in the foodservice industry. Sustainability; 14: 4893.

5. Canadian Centre for Occupational Health and Safety. (2022) Coronavirus (COVID-19) - tips: restaurants, bars, and food services (https://www.ccohs.ca/covid19/restaurants/). Accessed 3 January 2023.

6. Angelo KM, et al. (2017) Epidemiology of restaurant-associated foodborne disease outbreaks, United States, 1998–2013. Epidemiology and Infection; 145: 523–534.

7. Ontario Ministry of Health and Long-Term Care. (2019) Operational approaches for food safety guideline (https://www.health.gov.on.ca/en/pro/programs/publichealth/oph_standards/docs/protocols_guidelines/Operational_Approaches_For_Food_Safety_Guideline_2019_en.pdf). Accessed 3 January 2023.

8. City of Toronto. (2022) About DineSafe (https://www.toronto.ca/community-people/health-wellness-care/health-programs-advice/food-safety/dinesafe/about-dinesafe/). Accessed 3 January 2023.

9. Firestone MJ, et al. (2020) Can aggregated restaurant inspection data help us understand why individual foodborne illness outbreaks occur? Journal of Food Protection; 83: 788–793.

10. Fleetwood J, et al. (2019) As clean as they look? Food hygiene inspection scores, microbiological contamination, and foodborne illness. Food Control; 96: 76–86.

11. Petran RL, White BW, Hedberg CW. (2012) Health department inspection criteria more likely to be associated with outbreak restaurants in Minnesota. Journal of Food Protection; 75: 2007–2015.

12. City of Toronto. (2022) DineSafe (https://open.toronto.ca/dataset/dinesafe/). Accessed 3 January 2023.

13. R Core Team. (2022) R: a language and environment for statistical computing. Vienna, Austria: R Foundation for Statistical Computing.

14. RStudio Team. (2020) RStudio: integrated development for R. Boston, MA: RStudio, PBC.

15. Bhaskaran K, et al. (2013) Time series regression studies in environmental epidemiology. International Journal of Epidemiology; 42: 1187–1195.

16. Lopez Bernal J, et al. (2017) Interrupted time series regression for the evaluation of public health interventions: a tutorial. International Journal of Epidemiology; 46: 348–355.

17. McElreath R. (2020) Statistical Rethinking, 2nd ed. Boca Raton, FL: Chapman and Hall/CRC.

18. van de Schoot R, et al. (2021) Bayesian statistics and modelling. Nature Reviews Methods Primers; 1: 1–26.

19. Vehtari A, Gelman A, Gabry J. (2017) Practical Bayesian model evaluation using leave-one-out cross-validation and WAIC. Statistics and Computing; 27: 1413--1432.

20. Bürkner PC. (2017) brms: an R package for Bayesian multilevel models using Stan. Journal of Statistical Software; 80: 1–28.

21. Carpenter B, et al. (2017) Stan: a probabilistic programming language. Journal of Statistical Software; 76: 1–32.

22. Gabry J, ČeŠnovar R. (2022) cmdStanR (https://mc-stan.org/cmdstanr/index.html). Accessed 3 January 2023.

23. Gabry J, et al. (2019) Visualization in Bayesian workflow. Journal of the Royal Statistical Society: Series A; 182: 389–402.

24. Depaoli S, van de Schoot R. (2017) Improving transparency and replication in Bayesian statistics: the WAMBS-Checklist. Psychological Methods; 22: 240–261.

25. Arel-Bundock V. (2022) marginaleffects: marginal effects, marginal means, predictions, and contrasts (https://vincentarelbundock.github.io/marginaleffects/). Accessed 3 January 2023.

26. Sekercioglu F, et al. (2020) Experiences of environmental public health professionals during the COVID-19 pandemic response in Canada. Environmental Health Review; 63: 70–76.

27. Haas R, et al. (2021) ‘I walk around like my hands are covered in mud’: food safety and hand hygiene behaviors of Canadians during the COVID-19 pandemic. Food Protection Trends; 41: 454–463.

28. Thomas MS, Feng Y. (2021) Food handling practices in the era of COVID-19: a mixed-method longitudinal needs assessment of consumers in the United States. Journal of Food Protection; 84: 1176–1187.

29. Government of Ontario. (2022) Report on amendments, extensions, and revocations of orders under the Reopening Ontario (A Flexible Response to COVID-19) Act, 2020 from December 2, 2021, to March 28, 2022 (http://www.ontario.ca/document/report-amendments-extensions-and-revocations-orders-under-reopening-ontario-flexible-3). Accessed 3 January 2023.

30. Jeong M, et al. (2022) Key factors driving customers’ restaurant dining behavior during the COVID-19 pandemic. International Journal of Contemporary Hospitality Management; 34: 836–858.

31. Messabia N, Fomi P-R, Kooli C. (2022) Managing restaurants during the COVID-19 crisis: innovating to survive and prosper. Journal of Innovation and Knowledge; 7: 100234.

32. Dominianni C, et al. (2018) Hot weather impacts on New York City restaurant food safety violations and operations. Journal of Food Protection; 81: 1048–1054.

33. Powell MR, et al. (2018) Temporal patterns in principal Salmonella serotypes in the USA; 1996–2014. Epidemiology and Infection; 146: 437–441.

34. Simpson RB, Zhou B, Naumova EN. (2020) Seasonal synchronization of foodborne outbreaks in the United States, 1996–2017. Scientific Reports; 10: 17500.

35. Harris KJ, et al. (2015) Food safety inspections results: a comparison of ethnic-operated restaurants to non-ethnic-operated restaurants. International Journal of Hospitality Management; 46: 190–199.

36. Leinwand SE, et al. (2017) Inspection frequency, sociodemographic factors, and food safety violations in chain and nonchain restaurants, Philadelphia, Pennsylvania, 2013-2014. Public Health Reports; 132: 180–187.

37. Menachemi N, et al. (2012) Characteristics of restaurants associated with critical food safety violations. Food Protection Trends; 32: 73–80.

38. Ng DLT, et al. (2020) Spatial distribution and characteristics of restaurant inspection results in Toronto, Ontario, 2017–2018. Food Protection Trends; 40: 232–240.

