## Supplementary material for "Impact of the COVID-19 pandemic on food safety inspection outcomes in Toronto, Canada: a Bayesian interrupted time series analysis"

**Bayesian Regression Model Diagnostic Charts and Sensitivity Analysis**

**Pass rate model:**

LOO model comparison

| Model | ELPD difference | SE of difference |
| --- | --- | --- |
| Month as varying effect | 0 | 0 |
| Month as fixed-effect indicator | -1.0 | 1.6 |
| Month as varying effect with first-order autoregressive term | -3.2 | 4.6 |
| Month as fixed-effect indicator with first-order autoregressive term | -6.0 | 4.5 |

MCMC chains and posterior parameter distributions

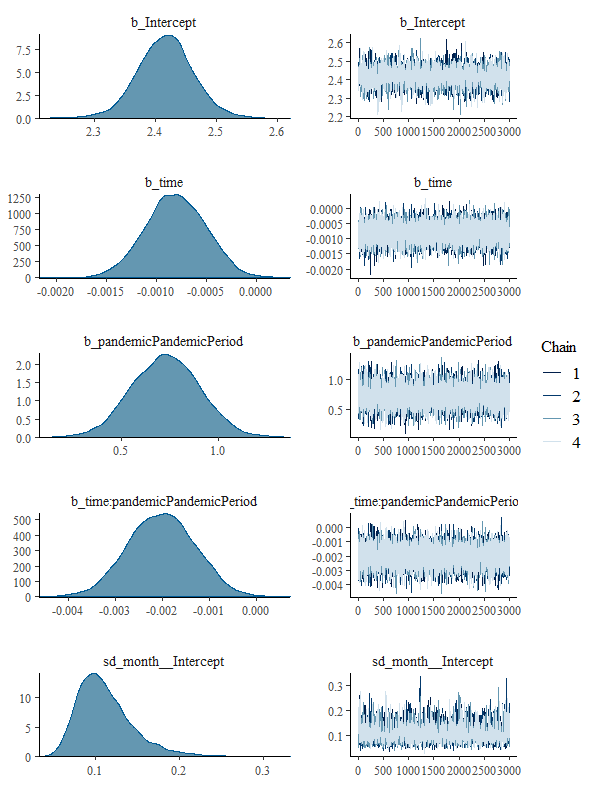

Posterior prediction check (100 draws)

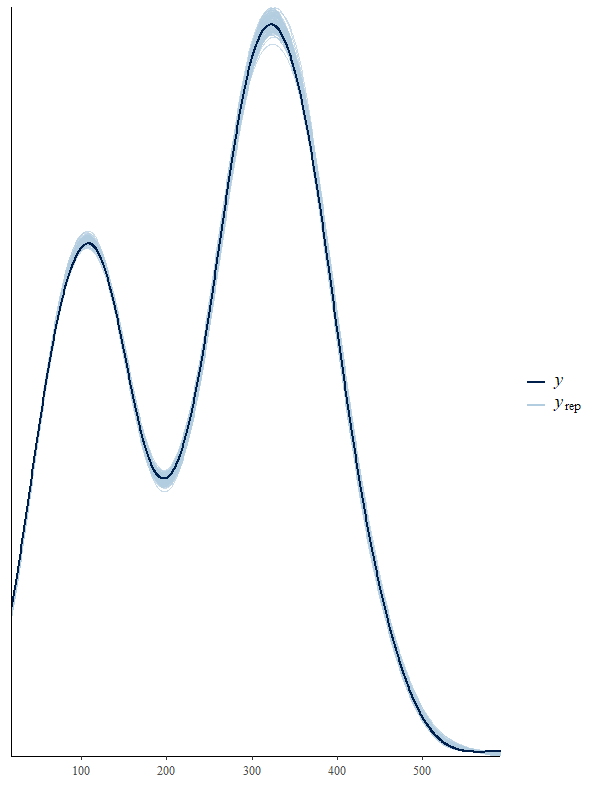

Residual autocorrelation for model parameters

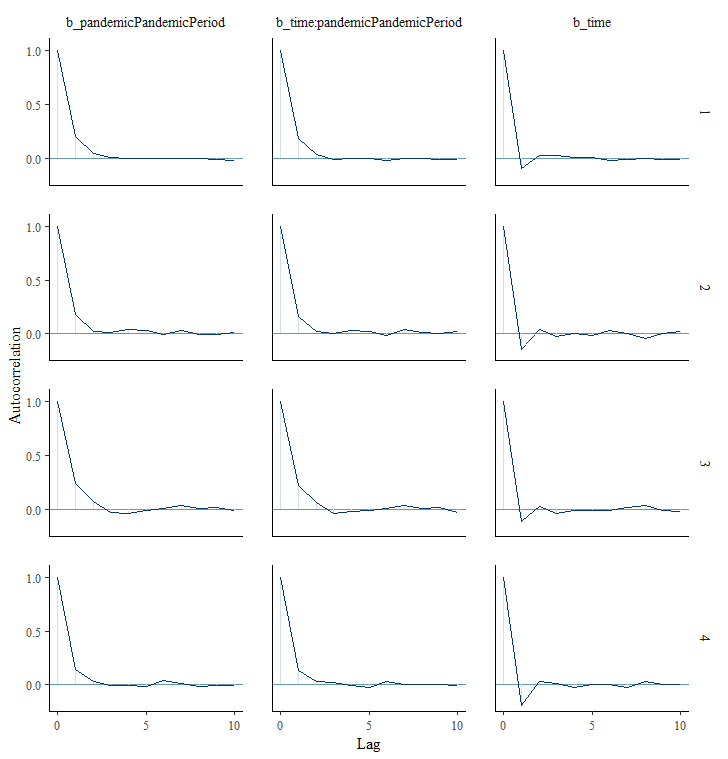

Sensitivity analysis of alternative prior distributions

|  | Normal(0, 1) prior | | Normal(0, 0.5) prior | | Normal(0, 2) prior | |
| --- | --- | --- | --- | --- | --- | --- |
| Parameter | Estimate | 95% CI | Estimate | 95% CI | Estimate | 95% CI |
| Intercept | 2.417 | 2.325, 2.508 | 2.403 | 2.314, 2.487 | 2.419 | 2.332, 2.505 |
| Time elapsed | -0.0008 | -0.0014,  -0.0002 | -0.0008 | -0.0014, -0.0002 | -0.0008 | -0.0014,  -0.0002 |
| Pandemic period (yes vs. no) | 0.731 | 0.390, 1.073 | 0.674 | 0.358, 0.999 | 0.746 | 0.408, 1.085 |
| Time*pandemic interaction term | -0.0020 | -0.0034,  -0.0006 | -0.0018 | -0.0032, -0.0005 | -0.0021 | -0.0035, -0.0007 |
| Group-level effects for month | 0.111 | 0.063, 0.191 | 0.111 | 0.063, 0.189 | 0.111 | 0.063, 0.188 |

**Infraction rate model:**

LOO model comparison

| Model | ELPD difference | SE of difference |
| --- | --- | --- |
| Month as varying effect | 0 | 0 |
| Month as fixed-effect indicator | -0.6 | 1.5 |
| Month as fixed-effect indicator with first-order autoregressive term | -48.6 | 10.3 |
| Month as varying effect with first-order autoregressive term | -51.2 | 10.3 |

MCMC chains and posterior parameter distributions

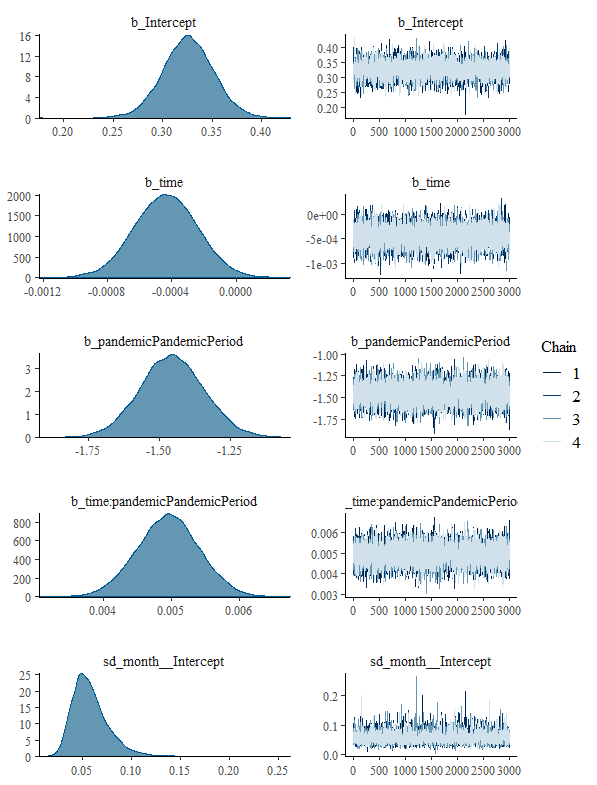

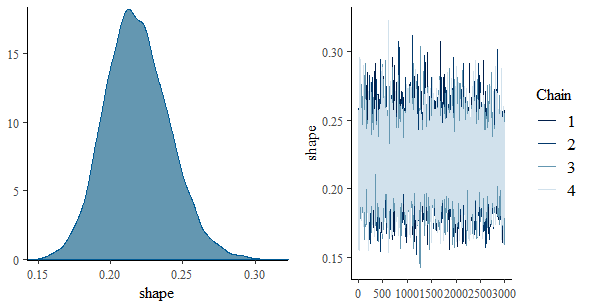

Posterior prediction check (100 draws)

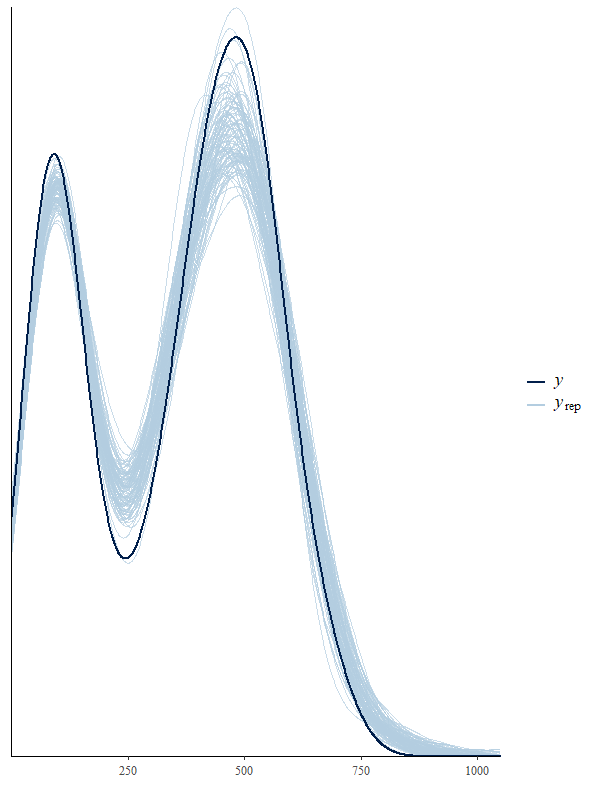

Residual autocorrelation for model parameters

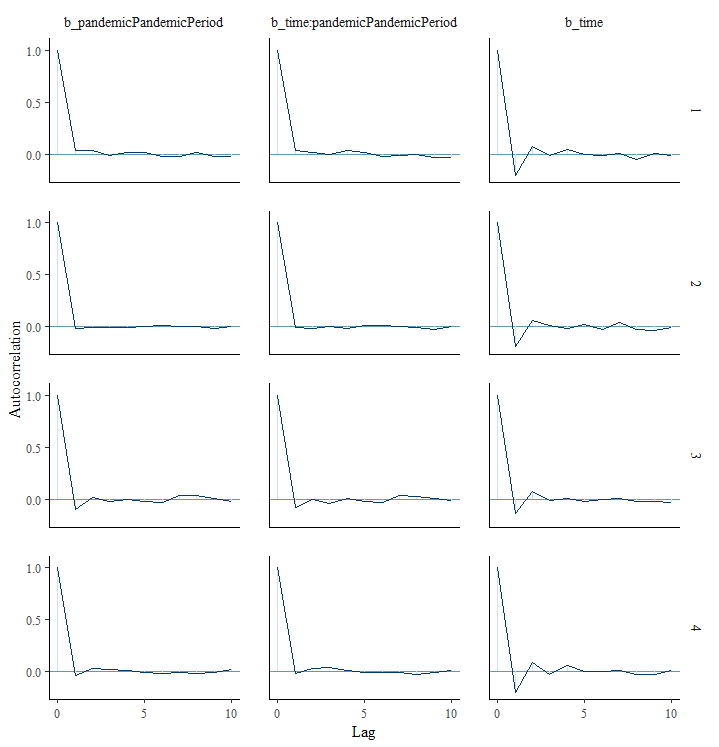

Sensitivity analysis of alternative prior distributions

|  | Normal(0, 1) prior | | Normal(0, 0.5) prior | | Normal(0, 2) prior | |
| --- | --- | --- | --- | --- | --- | --- |
| Parameter | Estimate | 95% CI | Estimate | 95% CI | Estimate | 95% CI |
| Intercept | 0.326 | 0.274, 0.377 | 0.322 | 0.269, 0.372 | 0.325 | 0.274, 0.376 |
| Time elapsed | -0.0004 | -0.0008, -0.0001 | -0.0004 | -0.0008, -0.00002 | -0.0004 | -0.0008, -0.0001 |
| Pandemic period (yes vs. no) | -1.456 | -1.673,  -1.235 | -1.400 | -1.616,  -1.178 | -1.468 | -1.686,  -1.248 |
| Time*pandemic interaction term | 0.005 | 0.004, 0.006 | 0.005 | 0.004, 0.006 | 0.005 | 0.004, 0.006 |
| Group-level effects for month | 0.058 | 0.030, 0.103 | 0.058 | 0.030, 0.100 | 0.058 | 0.030, 0.105 |
